# Risk Factors for Post-Injection Endophthalmitis: A Retrospective National Study in the IRIS^®^ Registry (Intelligent Research in Sight)

**DOI:** 10.64898/2026.01.12.25336174

**Authors:** Sophia Y. Ghauri, Connor Ross, Joshua B. Gilbert, Daniel J. Hu, Dan Gong, Paul B Greenberg, Dean Eliott, Tobias Elze, Alice Lorch, Joan W Miller, Magdalena G. Krzystolik, the IRIS® Registry Analytic Center Consortium

**Author notes:** Contributed Equally (Sophia Y. Ghauri, Connor Ross). Corresponding Author **Address for Reprints:** Magdalena G. Krzystolik, MD, Mass Eye and Ear, Providence and Plainville, Assistant Professor of Ophthalmology, Harvard Medical School.

## Abstract

**Purpose:** To investigate the epidemiology of post-injection endophthalmitis (PIE) and evaluate the association of sociodemographic and clinical factors with incidence, timing of onset, and presenting visual acuity (VA) using the IRIS^®^ Registry (Intelligent Research in Sight).

**Design:** Retrospective cohort study.

**Participants:** Patients with endophthalmitis after an intravitreal anti-VEGF injection in the IRIS^®^ Registry from 2016-2023.

**Methods:** Only the first anti-VEGF injection per eye was included. Exclusion criteria were cataract surgery during the study, intravitreal corticosteroids within 30 days prior to PIE, uveitis, or cystoid macular edema. Mean best VA was recorded within 100 days prior to anti-VEGF treatment and at the time of PIE. Regression modeling evaluated associations between endophthalmitis and sociodemographic and clinical factors, and time to PIE. Linear regression assessed predictors of VA at the time of PIE, and descriptive statistics were used to analyze time to onset.

**Main Outcome Measures:** Incidence of post-injection endophthalmitis, time to symptom onset, and best VA at diagnosis.

**Results:** Among 1,025,788 eyes treated, 600 (0.059%) developed endophthalmitis. Key risk factors included residence in U.S. territories (OR = 2.62; *P* = 0.038 vs. Northeast) and history of intravitreal corticosteroid injection (OR = 2.35; *P* = 0.004 vs. no history). The strongest protective factor was non-smoking (OR = 0.71; *P* = 0.023 vs. smokers). The median time from injection to onset of PIE was 5 days (interquartile range [IQR]: 3–8). The salient predictors of time to PIE included patient age (4.3 days sooner per decade older; *P* = 0.04), prior corticosteroid treatment (11.7 days sooner; *P* = 0.02), and a diagnosis of diabetic retinopathy (2.3 days sooner; *P*=0.03). Baseline VA before PIE was the only significant predictor of VA at the time of PIE diagnosis (0.67, *P* < 0.001).

**Conclusion:** Post-injection endophthalmitis was significantly associated with residence in U.S. territories and prior intravitreal corticosteroid exposure, while non-smoking status was protective. Most cases presented 3-8 days following anti-VEGF injection. Older age, history of prior corticosteroid treatment, and diabetic retinopathy were associated with earlier PIE. Baseline VA predicted VA at the time of PIE.

## Introduction

Intravitreal anti-vascular endothelial growth factor (anti-VEGF) injections have transformed the management and visual prognosis of patients with retinal diseases, including exudative age-related macular degeneration (AMD), diabetic retinopathy (DR), diabetic macular edema (DME), and retinal vein occlusions (RVO), among others. Since their introduction in 2005, the annual volume of anti-VEGF injections administered in the United States has steadily increased.^1^ However, each injection carries a rare but potentially vision-devastating risk of post-injection endophthalmitis (PIE) if not promptly treated, which may lead to profound visual loss or even enucleation.^2^

In the United States (US), commonly cited papers report approximately 7 million intravitreal injections administered each year.^2,3^ Based on reported rates of PIE ranging from 0.028% to 0.056%, this predicts an estimated 2000 to 4000 cases of endophthalmitis annually.^2,3^

Patients requiring frequent, ongoing intravitreal therapy face an even greater cumulative risk of infection.^2,3^ Numerous hypotheses exist regarding the factors that may influence the risk of PIE, but few studies have comprehensively investigated its specific risk factors.^4^ Prior investigations have also been limited by small sample sizes, reducing the statistical power to detect meaningful associations.^4^

The investigation of rare ocular complications, including postprocedural endophthalmitis, has been facilitated by large-scale national databases and clinical registries such as those maintained by the US Department of Veterans Affairs, the Centers for Medicare & Medicaid Services, and the American Academy of Ophthalmology (Academy) IRIS^®^ Registry (Intelligent Research in Sight).^5,6^ We used data from the IRIS Registry to (1) identify risk factors for PIE, and among patients who develop PIE to (2) characterize the time to infection, and (3) evaluate the influence of selected sociodemographic and clinical variables on presenting visual acuity (VA) at the time of diagnosis.

## Methods

### Study Design and Data Source

We conducted a retrospective cohort study using deidentified data from the IRIS Registry. The IRIS Registry is a centralized data repository and reporting tool used for research purposes. This does not constitute human subject research because data in the IRIS Registry is de-identified and the investigator does not have access to study identifiers. Therefore, institutional board review and informed consent are not required. This study adheres to the Declaration of Helsinki.

Available data included demographic characteristics, diagnostic codes, procedural codes, clinical measurements, and longitudinal follow-up. This study’s data were drawn from the IRIS Registry database version “Chicago,” frozen on April 21, 2023, and accessed on November 21, 2024. All data extraction and transformation processes were performed using PostgreSQL and R statistical software (version 4.3.3).

Multivariable regression models were employed to evaluate associations between sociodemographic and clinical factors and the risk of developing PIE. Multivariable logistic regression was used to estimate odds ratios for incident PIE, while multivariable linear regression assessed predictors of time to onset. For patients who developed PIE, linear regression was also applied to identify factors associated with presenting VA at diagnosis. All models included relevant covariates described above, and statistical significance was determined at an alpha level of 0.05.

### Identification of Anti-VEGF Injections and PIE

We identified anti-VEGF intravitreal injections administered between January 1, 2016, and April 21, 2023, using the following HCPCS J codes: J9035 (bevacizumab), J0178 (aflibercept), J2778 (ranibizumab), J0179 (brolucizumab), and J3590 (unclassified biologic agent).^1^ To minimize misclassification from data duplication and to account for time-varying clinical covariates, only the first anti-VEGF injection per eye was selected for our study population. Records that included the same-day CPT code for intravitreal injection (67028) in the same eye were retained to confirm procedural integrity.^7^

Potential clinical indications for anti-VEGF treatment were identified by extracting all ophthalmic diagnosis International Classification of Diseases (ICD) codes recorded from 2016 onward for patients who received at least one qualifying anti-VEGF injection **(Supplementary Table 1)**. Diagnoses were retained only if they (1) occurred prior to or on the date of the first injection, (2) had known laterality, and (3) matched the laterality of the eye receiving the injection. Patients with no qualifying pre-injection diagnosis were assigned an “unknown” indication category.^2^

To evaluate the relevant ocular history, we extracted procedural data using Current Procedural Terminology (CPT) codes for cataract surgery, intravitreal corticosteroid administration, and glaucoma-related procedures **(Supplementary Table 2)**. Only records with laterality matching the injected eye were included.

Endophthalmitis events were identified using ICD-10 diagnosis codes and defined as any qualifying diagnosis occurring within 1 to 28 days following the first anti-VEGF injection. To ensure accurate attribution, only the first qualifying episode of endophthalmitis per eye was included; duplicate and subsequent diagnoses were excluded.

### Exclusion Criteria

Patients were excluded if they underwent cataract surgery on or after the date of their first anti-VEGF injection, had a documented history of intermediate or posterior uveitis any time prior to the diagnosis of endophthalmitis, or were diagnosed with cystoid macular edema (CME) without a concurrent diagnosis of retinal vein occlusion (RVO). Additional exclusion criteria included receiving intravitreal corticosteroids on or after the date of the first anti-VEGF injection or having any prior diagnosis of endophthalmitis before the first anti-VEGF injection.

### Sociodemographic and Clinical Covariates

Demographic and clinical variables were extracted at the time of the first anti-VEGF injection. Variables included age, sex, race and ethnicity, geographic region, smoking status, and insurance type.

### Visual Acuity Data Collection

Although not used in the primary risk factor analysis, VA data were extracted for patients who developed endophthalmitis. For each included eye, first, a baseline VA was extracted, which was calculated using the mean VA for all VA measurements within 100 days prior to the first anti-VEGF injection. Second, the VA at the time of diagnosis was collected to determine the VA at PIE presentation. VA values were standardized using IRIS Registry best practices for Snellen-to-logMAR conversion **(Supplementary Table 3)**. All analyses were conducted at the eye level, with each record representing a unique eye receiving its first anti-VEGF injection during the study period.

## Results

### Study Population

Between 2016 and April 1, 2023, a total of 1,025,788 eyes that received anti-VEGF injections were included, among which 600 cases of post-injection endophthalmitis were identified following the first anti-VEGF injection, corresponding to an overall incidence of 0.059%. There were no patients who developed a case of bilateral endophthalmitis on their first injection. The demographic and clinical characteristics of the cohort are described in **(Supplementary Table 4)**.

### Risk Factors Associated with PIE

Several factors were significantly associated with the risk of developing PIE. Residence in U.S. territories (Guam, American Samoa, Northern Mariana Islands, U.S. Virgin Islands, and Puerto Rico) was associated with increased odds of PIE compared to the Northeast region (OR = 2.62; *P* = 0.038; 95% CI: 1.05–6.53). A prior history of intravitreal corticosteroid injections at any time before the first anti-VEGF injection was also associated with a higher risk (OR = 2.35; *P* = 0.004; 95% CI: 1.32–4.20). In contrast, non-smokers had significantly lower odds of developing PIE compared to smokers (OR = 0.71; *P* = 0.023; 95% CI: 0.53–0.95), and patients treated with bevacizumab had a reduced risk compared to those receiving aflibercept (OR = 0.82; *P* = 0.040; 95% CI: 0.68–0.99 **(Table 1)**.

### Endophthalmitis Temporal Presentation

The median time from precipitating injection to onset of PIE was 5 days (IQR: 3–8). **Supplementary Figure 1** illustrates the distribution of days to PIE following the most recent anti-VEGF injection.

### Time to presentation

Several clinical and demographic factors were associated with a shorter time from anti-VEGF injection to presentation of PIE. Older patients presented with PIE earlier than their younger counterparts (4.28 days sooner per decade above the mean; *P* = 0.04). Earlier onset was also observed in those with a history of intravitreal corticosteroid use prior to anti-VEGF therapy compared to those without a history (11.66 days sooner; *P* = 0.02). Additionally, patients with diabetic retinopathy presented earlier than those with AMD (2.32 days sooner; *P* = 0.03), as did individuals residing in the Southern United States compared to those in the Northeast (2.14 days sooner; *P* = 0.04). **(Supplementary Table 5)**.

### VA at Endophthalmitis Diagnosis

Baseline VA was the only significant predictor of VA at the time of PIE diagnosis (0.09; *P* < 0.001). No other sociodemographic or clinical variables were significantly associated with presenting VA while adjusting for baseline VA, including days from injection to presentation/PIE diagnosis. **(Supplementary Table 6)**.

## Discussion

### Study Population

In this large-scale analysis of over 1 million eyes receiving intravitreal anti-VEGF injections in the IRIS Registry between 2016 and 2023, the overall incidence of PIE was 0.059%, similar to previous reported rates of 0.049% to 0.056%.^8–12^ PIE remains rare, with an incidence of approximately 1 in 1700 injections. With an estimated 7 million injections administered each year, this predicts approximately 4,000 cases of endophthalmitis annually.^2,3^

### Risk Factors Associated with PIE

Geographic variation emerged as a significant factor, with patients residing in U.S. territories showing higher odds of developing PIE compared to those in the Northeast, whereas other regions were not clinically significant. Limited access to specialized care in some of these regions may contribute to the increased risk.^17^ This finding suggests the need for targeted quality initiatives in underserved regions and further research into system-level factors influencing safety.

A prior history of intravitreal corticosteroid injections (before anti-VEGF therapy) was associated with increased PIE risk. This aligns with prior evidence that chronic corticosteroid use may confer local immunosuppression, increasing susceptibility to infection.^13,14^ VanderBeek et al. reported an odds ratio of 6.92 for endophthalmitis in patients receiving corticosteroids compared with anti-VEGF agents. ^15^ Similarly, the SCORE study, which evaluated intravitreal triamcinolone for BRVO, reported a 0.725% incidence of infectious endophthalmitis higher than with anti-VEGF injections.^16^ Our study suggests the additive effect, but previous studies are on corticosteroids injected alone. Our study suggests an additive effect, while some previous studies examined corticosteroids alone.

Smoking status was a relevant factor, with non-smokers demonstrating a significantly lower risk of PIE compared to smokers. This finding was supported by reports that smoking impaired ocular surface immunity and promoted inflammation, thereby increasing susceptibility to intraocular infections.^17,18^ Our data show that smoking may be a modifiable patient risk factor, and counseling smokers about eye infection risks is warranted.

Variation in the risk of PIE was observed across different anti-VEGF agents. Bevacizumab was associated with a reduced risk of PIE compared to aflibercept. In previous studies, Clay et al. reported higher odds of endophthalmitis with aflibercept and ranibizumab compared to sterilely preloaded bevacizumab, and Dhoot et al. observed lower rates of PIE with bevacizumab and ranibizumab (vial and pre-filled) compared to aflibercept.^19,20^ These findings underscore the potential safety advantage of sterile preloading in intravitreal injection protocols ^21^

### Endophthalmitis Temporal Presentation

The median time from injection to symptom onset was 5 days, with some associations with socio-demographic and clinical characteristics affecting the time to presentation of PIE. This timing was consistent with prior studies, including one by Dong et al., which reported a mean onset of 4.5 days following injection.^22^

Older patients tended to present with PIE sooner, potentially reflecting an age-related decline in immune function that impaired the ability to mount an effective response to infection.

Similarly, patients with prior intravitreal corticosteroid use were more likely to experience earlier PIE, possibly due to chronic corticosteroids’ known immunosuppressive effects increasing infection risk. While direct evidence for intravitreal corticosteroids is limited, long-term use of oral corticosteroids, which have similar immunosuppressive effects, has been associated with an increased risk of infections.^23^ Patients with diabetic retinopathy were more likely to exhibit an earlier presentation of PIE. This could be attributed to the impaired immune responses characteristic of DR, which facilitated pathogen entry into the vitreous cavity and accelerated infection onset.^24,25^

Patients living in the southern United States were more likely to present earlier with PIE relative to those in the Northeast. This could reflect immediate access to healthcare services or regional differences in healthcare-seeking behavior. Another possible explanation could be seasonal variation; prior studies had reported increased rates of post-cataract surgery endophthalmitis during hot and humid months, which may similarly influence the incidence and timing of post-injection endophthalmitis ^26^

### Visual Acuity at Time of Endophthalmitis Diagnosis

Baseline visual acuity was the only predictor of VA at the time of PIE diagnosis.

## Limitations

As a retrospective registry analysis, our study had several limitations. The IRIS Registry database is an electronic health record database and relies on diagnostic and procedural codes used primarily for billing purposes. We were unable to verify the microbiological culture results. Some variables, such as exact injection technique (mask use, speculum use) or environmental factors, were unavailable. The interval between symptom onset and intravitreal antibiotic administration could not be determined. The inclusion of unspecified data, for example, a missing diagnostic indication for anti-VEGF therapy, may have introduced bias or masked true associations. Despite these limitations, the very large sample size lends confidence to the identified associations, especially for relatively common covariates.

Although post-injection endophthalmitis occurs in only ~1 in 1700 injections, its impact is substantial given the volume of anti-VEGF use. Risk varies by geography, prior corticosteroid exposure, smoking, and anti-VEGF agent.

## Supporting information

IRIS Consortium Author List

Precis

## Data Availability

All data referenced in this manuscript are derived from the American Academy of Ophthalmology IRIS Registry. The Registry is a de‑identified, HIPAA‑compliant clinical database comprising tens of millions of patient encounters and accessible to researchers via AAO-approved pathways.

---

**Supplementary Figure 1:**
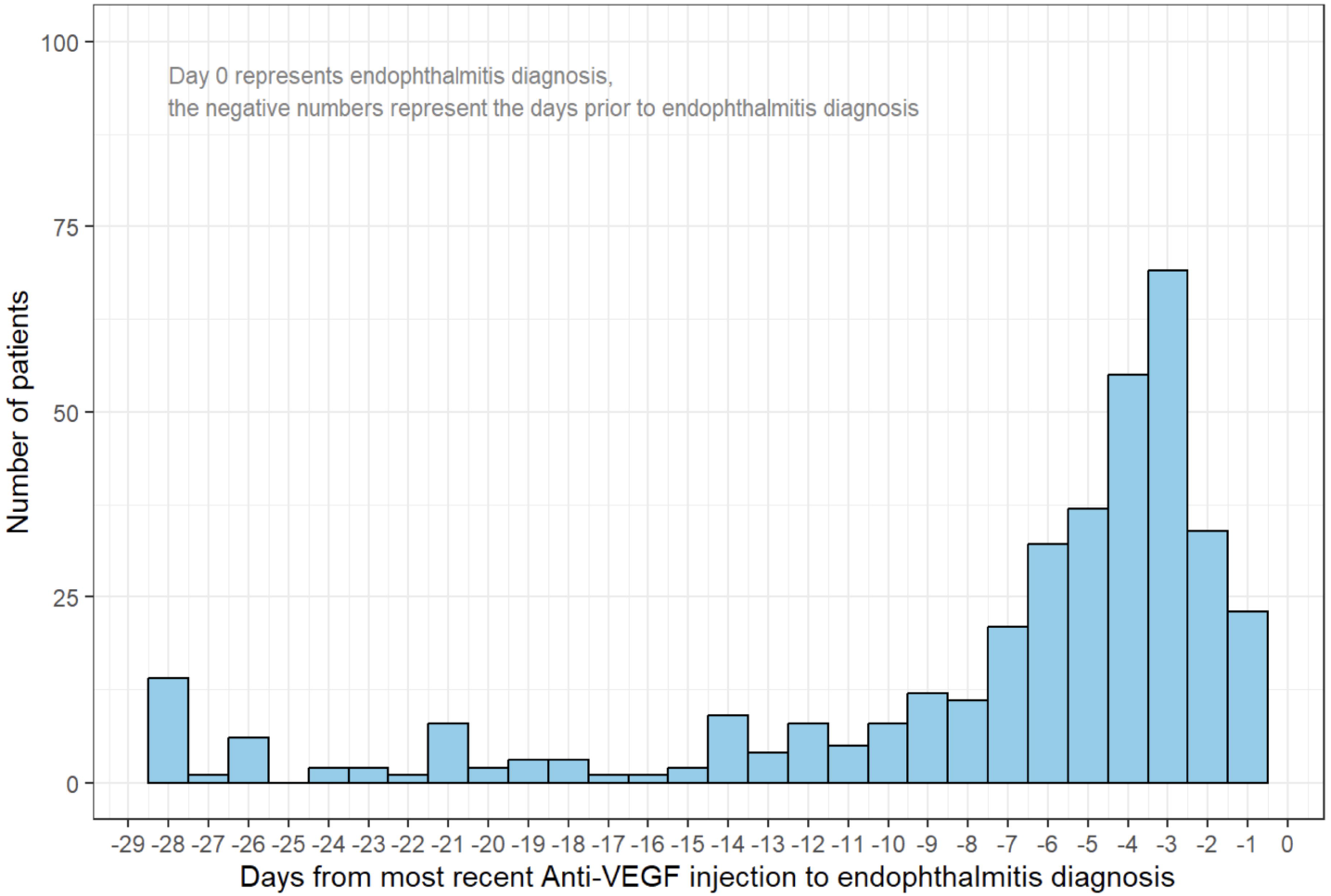
Days to PIE from Most Recent Anti-VEGF Injection.

## Footnotes

1 While newer agents were approved by the FDA in 2021 and 2022, Byooviz, Cimerli and Faricimab, there were no records of this treatment in IRIS’ 2023 version.

2 The IRIS Registry does not explicitly link diagnoses to procedures. As such, the indication for anti-VEGF therapy could not be directly determined and was instead inferred. We classified the indication based on the most recent FDA-approved condition for which anti-VEGF therapy is commonly applied. If two eligible diagnoses were recorded on the same day prior to injection, we imputed the indication as unknown.

## References

1. Israilevich RN, Mansour H, Patel SN, et al. Risk of Endophthalmitis Based on Cumulative Number of Anti-VEGF Intravitreal Injections. Ophthalmology 2024;131:667–673.

2. Patel SN, Gangaputra S, Sternberg P, Kim SJ. Prophylaxis measures for postinjection endophthalmitis. Surv Ophthalmol 2020;65:408–420.

3. Martin DF. Evolution of Intravitreal Therapy for Retinal Diseases—From CMV to CNV: The LXXIV Edward Jackson Memorial Lecture. Am J Ophthalmol 2018;191:xli–lviii.

4. Mccannel CA. META-ANALYSIS OF ENDOPHTHALMITIS AFTER INTRAVITREAL INJECTION OF ANTI–VASCULAR ENDOTHELIAL GROWTH FACTOR AGENTS: Causative Organisms and Possible Prevention Strategies. Retina 2011;31:654–661.

5. Lacy M, Kung T-PH, Owen JP, et al. Endophthalmitis Rate in Immediately Sequential versus Delayed Sequential Bilateral Cataract Surgery within the Intelligent Research in Sight (IRIS®) Registry Data. Ophthalmology 2022;129:129–138.

6. Lee CS, Blazes M, Lorch A, et al. American Academy of Ophthalmology Intelligent Research in Sight (IRIS®) Registry and the IRIS Registry Analytic Center Consortium. Ophthalmol Sci 2022;2:100112.

7. Goldberg E, Douglas V, Ivanov A, et al. Data Duplication and Errors in Large Medical Datasets: A Case Study in the IRIS® Registry. 2025. Available at: https://osf.io/gcqkt_v3 [Accessed July 31, 2025].

8. Mccannel CA. META-ANALYSIS OF ENDOPHTHALMITIS AFTER INTRAVITREAL INJECTION OF ANTI–VASCULAR ENDOTHELIAL GROWTH FACTOR AGENTS: Causative Organisms and Possible Prevention Strategies. Retina 2011;31:654–661.

9. Fileta JB, Scott IU, Flynn HW. Meta-Analysis of Infectious Endophthalmitis After Intravitreal Injection of Anti-Vascular Endothelial Growth Factor Agents. Ophthalmic Surg Lasers Imaging Retina 2014;45:143–149.

10. Storey P, Dollin M, Pitcher J, et al. The Role of Topical Antibiotic Prophylaxis to Prevent Endophthalmitis after Intravitreal Injection. Ophthalmology 2014;121:283–289.

11. Englander M, Chen TC, Paschalis EI, et al. Intravitreal injections at the Massachusetts Eye and Ear Infirmary: analysis of treatment indications and postinjection endophthalmitis rates. Br J Ophthalmol 2013;97:460.

12. Dossarps D, Bron AM, Koehrer P, et al. Endophthalmitis After Intravitreal Injections: Incidence, Presentation, Management, and Visual Outcome. Am J Ophthalmol 2015;160:17–25.e1.

13. Baudin F, Benzenine E, Mariet A-S, et al. Association of Acute Endophthalmitis With Intravitreal Injections of Corticosteroids or Anti–Vascular Growth Factor Agents in a Nationwide Study in France. JAMA Ophthalmol 2018;136:1352.

14. Hata M, Hata M, Dejda A, et al. Corticosteroids reduce pathological angiogenesis yet compromise reparative vascular remodeling in a model of retinopathy. Proc Natl Acad Sci U S A 121:e2411640121.

15. VanderBeek BL, Bonaffini SG, Ma L. The Association between Intravitreal Steroids and Post-Injection Endophthalmitis Rates. Ophthalmology 2015;122:2311–2315.e1.

16. Scott IU, Ip MS, VanVeldhuisen PC, et al. A Randomized Trial Comparing the Efficacy and Safety of Intravitreal Triamcinolone With Standard Care to Treat Vision Loss Associated With Macular Edema Secondary to Branch Retinal Vein Occlusion. Arch Ophthalmol 2009;127:1115–1128.

17. Galor A, Feuer W, Kempen JH, et al. Adverse effects of smoking on patients with ocular inflammation. Br J Ophthalmol 2010;94:848–853.

18. Anam A, Yu M, Liu C, et al. Smoking negatively impacts ocular surface health and corneal nerve metrics. Ocul Surf 2025;37:105–114.

19. Dhoot DS, Boucher N, Pitcher JD, Saroj N. Rates of Suspected Endophthalmitis Following Intravitreal Injections in Clinical Practices in the United States. Ophthalmic Surg Lasers Imaging Retina 2021;52:312–318.

20. Bavinger JC, Yu Y, VanderBeek BL. COMPARATIVE RISK OF ENDOPHTHALMITIS AFTER INTRAVITREAL INJECTION WITH BEVACIZUMAB, AFLIBERCEPT, AND RANIBIZUMAB. Retina 2019;39:2004–2011.

21. Brown DM, Sobel RE, Suzart-Woischnik K, et al. Intraocular Inflammation after Aflibercept Prefilled Syringe and Vial Injections. Ophthalmol Retina 2025;9:712–715.

22. Dong LK, Shields RA. Predictors of visual recovery after post-injection endophthalmitis. Invest Ophthalmol Vis Sci 2021;62:1350–1350.

23. Mustafa SS. Steroid-induced secondary immune deficiency. Ann Allergy Asthma Immunol 2023;130:713–717.

24. Xu H, Chen M. Immune response in retinal degenerative diseases – Time to rethink? Prog Neurobiol 2022;219:102350.

25. Semeraro F, Cancarini A, dell’Omo R, et al. Diabetic Retinopathy: Vascular and Inflammatory Disease. J Diabetes Res 2015;2015:582060.

26. Kim SH, Yu MH, Lee JH, et al. Seasonal variation in acute post-cataract surgery endophthalmitis incidences in South Korea. J Cataract Refract Surg 2019;45:1711–1716.

