## Supplemental Table 3 for "Risk Factors for Post-Injection Endophthalmitis: A Retrospective National Study in the IRIS^®^ Registry (Intelligent Research in Sight)"

**Supplementary Table 3: Snellen to LogMAR Conversion**

| ***Snellen Value*** | ***Calculated LogMAR*** |
| --- | --- |
| **20/10** | **-0.3** |
| **20/12.5** | **-0.2** |
| **20/15** | **-0.12** |
| **20/16** | **-0.1** |
| **20/20** | **0** |
| **20/25** | **0.1** |
| **20/30** | **0.18** |
| **20/32** | **0.2** |
| **20/40** | **0.3** |
| **20/50** | **0.4** |
| **20/60** | **0.48** |
| **20/63** | **0.5** |
| **20/65** | **0.51** |
| **20/70** | **0.54** |
| **20/80** | **0.6** |
| **20/100** | **0.7** |
| **20/120** | **0.78** |
| **20/125** | **0.8** |
| **20/150** | **0.88** |
| **20/160** | **0.9** |
| **20/200** | **1** |
| **20/250** | **1.1** |
| **20/300** | **1.18** |
| **20/320** | **1.2** |
| **20/350** | **1.24** |
| **20/400** | **1.3** |
| **20/500** | **1.4** |
| **20/600** | **1.48** |
| **20/630** | **1.5** |
| **20/650** | **1.51** |
| **20/800** | **1.6** |
| **5/200** | **1.6** |
| **20/1000** | **1.7** |
| **4/200** | **1.7** |
| **20/1200** | **1.78** |
| **20/1260** | **1.8** |
| **3/200** | **1.82** |
| **20/1600** | **1.9** |
| **5/400** | **1.9** |
| **CF** | **1.9** |
| **2/200** | **2** |
| **20/2000** | **2** |
| **4/400** | **2** |
| **3/400** | **2.12** |
| **1/200** | **2.3** |
| **2/400** | **2.3** |
| **HM** | **2.3** |
| **1/400** | **2.6** |
| **LP** | **2.7** |
| **NLP** | **4** |
