## Supplementary Table 4 for "Risk Factors for Post-Injection Endophthalmitis: A Retrospective National Study in the IRIS^®^ Registry (Intelligent Research in Sight)"

**Supplementary Table 4: Included Cohort Demographics**

| Groups | Non-PIE Group  N = 1,024,588 | PIE Group  N = 600 |
| --- | --- | --- |
| Anti-VEGF Drug |  |  |
| aflibercept | 308,389 (30%) | 198 (33%) |
| bevacizumab | 502,307 (49%) | 260 (43%) |
| brolucizumab | 1,688 (0.2%) | 2 (0.3%) |
| other | 72,251 (7.1%) | 57 (9.5%) |
| ranibizumab | 139,953 (14%) | 83 (14%) |
| Diagnosis |  |  |
| AMD | 412,419 (40%) | 217 (36%) |
| Degenerative Myopia with CNV | 4,261 (0.4%) | 1 (0.2%) |
| Diabetic Retinopathy | 223,996 (22%) | 103 (17%) |
| RVO with Cystoid Edema | 10,278 (1.0%) | 10 (1.7%) |
| Retinal Vein Occlusion | 75,278 (7.3%) | 47 (7.8%) |
| Unknown | 298,356 (29%) | 222 (37%) |
| Cataract Surgery or Lens Removal | 154,910 (15%) | 101 (17%) |
| History of Intravitreal Corticosteroid Use | 7,857 (0.8%) | 12 (2.0%) |
| Any Glaucoma Procedure | 9,049 (0.9%) | 9 (1.5%) |
| Age at anti-VEGF Injection | 75 (64, 83) | 75 (65, 84) |
| Unknown | 841 | 0 |
| Sex |  |  |
| Female | 573,677 (56%) | 330 (55%) |
| Male | 444,761 (43%) | 265 (44%) |
| Not Reported | 6,150 (0.6%) | 5 (0.8%) |
| Race |  |  |
| American Indian or Alaskan Native | 4,075 (0.4%) | 1 (0.2%) |
| Asian | 24,413 (2.4%) | 13 (2.2%) |
| Black or African American | 69,789 (6.8%) | 29 (4.8%) |
| Native Hawaiian or Other Pacific Islander | 1,859 (0.2%) | 1 (0.2%) |
| Other | 19,247 (1.9%) | 11 (1.8%) |
| Unknown | 153,302 (15%) | 85 (14%) |
| White | 751,903 (73%) | 460 (77%) |
| Ethnicity |  |  |
| Hispanic or Latino | 89,631 (8.7%) | 53 (8.8%) |
| Not Hispanic or Latino | 714,001 (70%) | 416 (69%) |
| Unknown | 220,956 (22%) | 131 (22%) |
| Region |  |  |
| Northeast | 200,527 (20%) | 108 (18%) |
| American Territory | 4,283 (0.4%) | 5 (0.8%) |
| North Central | 221,130 (22%) | 127 (21%) |
| South | 403,067 (40%) | 248 (41%) |
| West | 190,062 (19%) | 111 (19%) |
| Unknown | 5,519 | 1 |
| Smoking Status |  |  |
| Active | 72,925 (7.1%) | 54 (9.0%) |
| Former | 217,711 (21%) | 125 (21%) |
| Never | 560,896 (55%) | 305 (51%) |
| Unknown | 173,056 (17%) | 116 (19%) |
| Insurance Status |  |  |
| Commercial | 144,469 (14%) | 90 (15%) |
| Govt | 7,032 (0.7%) | 2 (0.3%) |
| Medicare | 201,158 (20%) | 130 (22%) |
| Military | 5,368 (0.5%) | 2 (0.3%) |
| Multiple insurances listed | 103,048 (10%) | 61 (10%) |
| No insurance | 2,084 (0.2%) | 0 (0%) |
| Unknown | 561,429 (55%) | 315 (53%) |
| n (%); Median (Q1, Q3) | | |
