## Supplementary Table 5 for "Risk Factors for Post-Injection Endophthalmitis: A Retrospective National Study in the IRIS^®^ Registry (Intelligent Research in Sight)"

**Supplementary Table 5: Sociodemographic and Clinical Risk Factors Associated with Time to PIE Onset**

| Variable | Estimate | Standard Error | P-value |
| --- | --- | --- | --- |
| (Intercept) | 4.1 | 3.92 | 0.30 |
| Bevacizumab Treatment | 0.6 | 0.86 | 0.51 |
| Brolucizumab Treatment | 5.6 | 4.86 | 0.25 |
| **Unknown Anti-VEGF Treatment*** | 3.3 | 1.4 | 0.02 |
| Ranibizumab Treatment | 0.4 | 1.27 | 0.75 |
| **Degenerative Myopia with CNV*** | -3.2 | 6.86 | 0.65 |
| Diabetic Retinopathy | 2.3 | 1.07 | 0.03 |
| RVO with Cystoid Edema | 5.9 | 3.08 | 0.06 |
| Retinal Vein Occlusion | 0.0 | 1.37 | 0.99 |
| Diagnosis Unknown† | 2.7 | 0.9 | 0.00 |
| Prior GP Trabeculectomy | -6.8 | 9.78 | 0.49 |
| Prior GP Tube Shunt | 4.9 | 8.51 | 0.56 |
| No Prior Treatment | 0.3 | 0.96 | 0.76 |
| Prior Corticosteroid* | 11.7 | 4.89 | 0.02 |
| **Patient Age (per 10 Years)*** | 4.3 | 2.11 | 0.04 |
| Male Sex | 0.2 | 0.75 | 0.79 |
| Sex Not Reported | -1.0 | 3.07 | 0.76 |
| American Indian or Alaska Native | -6.1 | 7.32 | 0.40 |
| Black or African American | -4.7 | 3.02 | 0.12 |
| Other Race | 0.3 | 3.92 | 0.93 |
| Race Unknown | -0.8 | 2.88 | 0.79 |
| White | -2.3 | 2.63 | 0.39 |
| Not Hispanic or Latino | 0.8 | 1.37 | 0.56 |
| Ethnicity Unknown | 2.1 | 1.58 | 0.19 |
| American Territory | 5.2 | 6.91 | 0.45 |
| Region: North Central | -0.2 | 1.16 | 0.84 |
| **Region: South*** | 2.1 | 1.05 | 0.04 |
| Region: West | 0.1 | 1.2 | 0.94 |
| Former Smoker | -1.2 | 1.36 | 0.38 |
| Never Smoked | -1.4 | 1.2 | 0.26 |
| Smoking Status Unknown | -0.8 | 1.38 | 0.58 |
| Medicare Insurance | -0.1 | 1.21 | 0.95 |
| Military Insurance | 0.3 | 4.99 | 0.95 |
| Multiple Insurance Types | -0.8 | 1.43 | 0.59 |
| Insurance Status Unknown | -0.5 | 1.05 | 0.62 |
| Any Prior Glaucoma Procedure | 2.7 | 7.06 | 0.70 |
| Cataract Surgery or Lens Removal | NA | NA | NA |
| Corticosteroid Treatment | NA | NA | NA |
| **OR** = odds ratio, **LB** = lower bound, **UB** = upper bound | | N = 600  R^2^ = 0.031  R^2^ adjusted = 0.024  * P < 0.05  † P < 0.001 |  |
