## Supplementary Table 6 for "Risk Factors for Post-Injection Endophthalmitis: A Retrospective National Study in the IRIS^®^ Registry (Intelligent Research in Sight)"

**Supplementary Table 6: Sociodemographic and Clinical Risk Factors Associated with VA at PIE Diagnosis**

| Variable | Estimate | Standard Error | P-value |
| --- | --- | --- | --- |
| (Intercept) | 1.03 | 1.03 | 0.32 |
| **Mean LogMAR Before Anti-VEGF†** | 0.67 | 0.09 | 0.00 |
| Degenerative Myopia with CNV | 0.4 | 0.81 | 0.62 |
| Diabetic Retinopathy | 0.07 | 0.17 | 0.70 |
| RVO with Cystoid Edema | -0.51 | 0.3 | 0.10 |
| Retinal Vein Occlusion | -0.22 | 0.19 | 0.24 |
| Diagnosis Unknown | -0.05 | 0.13 | 0.72 |
| Prior GP MIGS | -0.26 | 0.92 | 0.78 |
| Prior GP Tube Shunt | -0.17 | 1 | 0.87 |
| No Prior Treatment | -0.36 | 0.86 | 0.67 |
| Prior Corticosteroid | -0.16 | 0.82 | 0.84 |
| Patient Age (per 10 Years) | 0 | 0 | 0.41 |
| Male Sex | 0.02 | 0.1 | 0.83 |
| Sex Not Reported | -0.48 | 0.81 | 0.55 |
| Black or African American | 0.3 | 0.48 | 0.54 |
| Native Hawaiian or Other Pacific Islander | -0.04 | 0.9 | 0.97 |
| Other Race | -0.13 | 0.64 | 0.84 |
| Race Unknown | 0.31 | 0.45 | 0.49 |
| White | 0.27 | 0.41 | 0.51 |
| Former Smoker | 0.09 | 0.2 | 0.64 |
| Never Smoked | -0.05 | 0.18 | 0.79 |
| Smoking Status Unknown | 0.07 | 0.2 | 0.71 |
| Government Insurance | -0.61 | 0.82 | 0.46 |
| Medicare Insurance | 0 | 0.16 | 1.00 |
| Military Insurance | -0.73 | 0.86 | 0.39 |
| Multiple Insurance Types | 0.09 | 0.19 | 0.65 |
| Insurance Status Unknown | -0.12 | 0.14 | 0.40 |
| Cataract Surgery or Lens Removal | -0.3 | 0.85 | 0.72 |
| Days from Diagnosis to Anti-VEGF | 0.01 | 0.01 | 0.30 |
| Any Prior Glaucoma Procedure | NA | NA | NA |
| Corticosteroid Treatment | NA | NA | NA |
| **OR** = odds ratio, **LB** = lower bound, **UB** = upper bound | | N = 600  R^2^ = 0.213  R^2^ adjusted = 0.205  * P < 0.05  † P < 0.001 | |
