## Supplementary material for "Risk Factors for Post-Injection Endophthalmitis: A Retrospective National Study in the IRIS^®^ Registry (Intelligent Research in Sight)": Table 1

**Table 1: Sociodemographic and Clinical Risk Factors Associated with PIE**

| Variable | Estimate | P-value | OR | LB | UB |
| --- | --- | --- | --- | --- | --- |
| **(Intercept)** | -8.33 | 0.000 | 0.00 | 0.00 | 0.00 |
| Patient Age | 0.00 | 0.094 | 1.00 | 1.00 | 1.00 |
| **Sex (Ref: Female)** | | | | | |
| Male Sex | 0.03 | 0.727 | 1.03 | 0.87 | 1.22 |
| Sex Not Reported | 0.37 | 0.411 | 1.45 | 0.60 | 3.51 |
| **Race (Ref: Asian)** | | | | | |
| American Indian or Alaska Native | -0.81 | 0.438 | 0.45 | 0.06 | 3.42 |
| Black or African American | -0.30 | 0.373 | 0.74 | 0.38 | 1.44 |
| Native Hawaiian or Other Pacific Islander | 0.03 | 0.979 | 1.03 | 0.13 | 7.88 |
| Other Race | 0.03 | 0.955 | 1.03 | 0.43 | 2.43 |
| Unknown Race | -0.07 | 0.834 | 0.94 | 0.51 | 1.72 |
| White | 0.06 | 0.826 | 1.07 | 0.61 | 1.86 |
| **Ethnicity (Ref: Hispanic or Latino)** | | | | | |
| Not Hispanic or Latino | -0.09 | 0.591 | 0.91 | 0.65 | 1.27 |
| Unknown Ethnicity | 0.04 | 0.833 | 1.04 | 0.72 | 1.50 |
| **Region (Ref: Northeast)** | | | | | |
| **Region: American Territory*^** | **0.96** | **0.038** | **2.62** | **1.05** | **6.53** |
| Region: North Central | 0.02 | 0.888 | 1.02 | 0.78 | 1.33 |
| Region: South | 0.17 | 0.144 | 1.19 | 0.94 | 1.49 |
| Region: West | 0.13 | 0.354 | 1.14 | 0.87 | 1.49 |
| **Smoking Status (Ref: Active smoker)** | | | | | |
| Former Smoker | -0.30 | 0.066 | 0.74 | 0.53 | 1.02 |
| **Never Smoked*** | **-0.34** | **0.023** | **0.71** | **0.53** | **0.95** |
| Smoking Status Unknown | -0.17 | 0.322 | 0.85 | 0.61 | 1.18 |
| **Insurance Status (Ref: Commercial insurance)** | | | | | |
| Government Insurance | -0.83 | 0.249 | 0.44 | 0.11 | 1.78 |
| Medicare Insurance | -0.03 | 0.845 | 0.97 | 0.74 | 1.28 |
| Military Insurance | -0.61 | 0.394 | 0.54 | 0.13 | 2.21 |
| Multiple Insurances Listed | -0.12 | 0.494 | 0.89 | 0.64 | 1.24 |
| No Insurance | -11.15 | 0.938 | 0.00 | 0.00 | 23.40 |
| Insurance Status Unknown | -0.11 | 0.367 | 0.90 | 0.71 | 1.14 |
| **Anti-VEGF Agent injected (Ref: Aflibercept)** | | | | | |
| **Bevacizumab Treatment*** | **-0.20** | **0.040** | **0.82** | **0.68** | **0.99** |
| Brolucizumab Treatment | 0.62 | 0.384 | 1.86 | 0.46 | 7.52 |
| Other Anti-VEGF Treatment | 0.25 | 0.112 | 1.29 | 0.94 | 1.75 |
| Ranibizumab Treatment | -0.06 | 0.634 | 0.94 | 0.73 | 1.22 |
| **Clinical History (Ref: No indication)** | | | | | |
| Cataract Surgery or Lens Removal | 0.07 | 0.521 | 1.08 | 0.86 | 1.35 |
| **Corticosteroid Treatment*** | **0.86** | **0.004** | **2.35** | **1.32** | **4.20** |
| History of Any Glaucoma Procedure | 0.46 | 0.180 | 1.58 | 0.81 | 3.09 |
| **Indication for Anti-VEGF (Ref: exudative AMD)** | | | | | |
| Degenerative Myopia with Choroidal Neovascularization | -0.62 | 0.540 | 0.54 | 0.08 | 3.88 |
| Diabetic Retinopathy (including DME) | 0.02 | 0.912 | 1.02 | 0.78 | 1.32 |
| Retinal Vein Occlusion with Cystoid Macular Edema | 0.59 | 0.070 | 1.81 | 0.95 | 3.43 |
| Retinal Vein Occlusion | 0.24 | 0.140 | 1.27 | 0.92 | 1.75 |
| **Unknown Diagnosis*** | **0.42** | **0.000** | **1.52** | **1.25** | **1.86** |
| **OR** = odds ratio, **LB** = lower bound, **UB** = upper bound  **Ref** = Reference Group | N = 1,025,788  * P < 0.05  † P < 0.001  ^American Territory Refers to: Guam, American Samoa, Northern Mariana Islands, District of Columbia, U.S. Virgin Islands, Puerto Rico | | | | |
