## Supplementary material for "Risk Factors for Post-Injection Endophthalmitis: A Retrospective National Study in the IRIS^®^ Registry (Intelligent Research in Sight)": IRIS Consortium Author List

IRIS Consortium Author List:
Nisha Acharya, MD, MS

Emily Y Chew, MD

Julia A Haller, MD

Leslie G Hyman, PhD

Aaron Y Lee, MD, MSCI

Cecilia S Lee, MD, MS

Alice C Lorch, MD

Flora Lum, MD

Felipe Andrade Medeiros, MD, PhD

Joan W Miller, MD

Alexander Roland Miranda, MD

Suzann Pershing, MD

Divya S Srikumaran, MD

Catherine Q Sun, MD

Swarup S Swaminathan, MD

Christina L Thomas-Virnig, PhD

Fasika A Woreta, MD, MPH
