## Supplementary material for "Risk Factors for Post-Injection Endophthalmitis: A Retrospective National Study in the IRIS^®^ Registry (Intelligent Research in Sight)": Precis

Precis (35-words):
This IRIS Registry study found post-injection endophthalmitis risk increased with residence in U.S. territories, smoking, and prior corticosteroids; bevacizumab was associated with lower risk. Most cases presented 3–8 days after anti-VEGF injection.
