## Supplementary Table 1 for "Risk Factors for Post-Injection Endophthalmitis: A Retrospective National Study in the IRIS^®^ Registry (Intelligent Research in Sight)"

**Supplementary Table 1: ICD Codes Included**

| ***Condition*** | ***ICD-10 Codes^1^*** |
| --- | --- |
| **Endophthalmitis** | H44.00%, H44.19% |
| **Uveitis** | H30.2% |
| **Cystoid Macular Edema (CME)** | H59.03% |
| **Diabetic Retinopathy** | E08.3%, E09.3%, E10.3%, E11.3%, E13.3% |
| **Exudative Age-Related Macular Degeneration** | H35.32% |
| **Retinal Vein Occlusion** | E34.81%, H34.83% |
| **Cystoid Macular Edema** | H35.35%, H59.03% |
| **Choroidal Neovascularization** | H35.05% |
| **Degenerative Myopia + CNV** | H44.2A% |
| **Retinal Neovascularization** | H35.059 |

**^1^** *The percent sign (%) denotes a wildcard of any number of alphanumeric characters*
