## Supplementary Table 2 for "Risk Factors for Post-Injection Endophthalmitis: A Retrospective National Study in the IRIS^®^ Registry (Intelligent Research in Sight)"

**Supplementary Table 2: CPT Codes Included**

| ***Procedure Category*** | ***Procedure Description*** | ***CPT Codes*** |
| --- | --- | --- |
| **Glaucoma Procedure** | **MIGS** | 0449T, 0253T, 65820, 0191T, 65850, 0450T, 0474T, 66183, 0376T |
|  | **MIGS with Cataract Surgery** | 66989, 66991, 0671T, 0191T, 0376T |
|  | **Tube Shunts** | 66180, 66179, 66183, 66184, 66185, 65920, 0192T |
|  | **Trabeculectomy** | 66172, J7315, 67255, 66150, 66155, 66160, 66170, 66250, 66165 |
|  | **Other Glaucoma Surgery** | 66250, 65820, 66174, 0177T, 66500, 66990, 66175, 0176T, 66505, 66600, 66605, 66625, 66630, 66635 |
| **Anti-VEGF** | **Aflibercept** | J0178 |
|  | **Brolucizumab** | J0179 |
|  | **Bevacizumab** | J9035 |
|  | **Ranibizumab** | J2778 |
|  | **Other** | J3590 |
| **Lens Removal** | **Cataract surgery or lens removal** | 66840, 66850, 66852, 66920, 66930, 66940, 66983, 66984, 66988, 66982, 66987 |
| **Corticosteroid** | **Steroid Injection** | J3300, J3301 |
